# Mind the Gap: Unraveling Mental Health Disparities in America’s Diverse Landscape

**DOI:** 10.1101/2024.07.28.24311109

**Authors:** Margaret Fischer, Jennifer Swint, Wei Zhang, Xi Zhang

## Abstract

**Background:** Mental health disparities persist as a significant public health concern in the United States, with certain populations experiencing disproportionate burdens of mental illness and barriers to care. This systematic review aims to synthesize current evidence on mental health disparities across various demographic groups and identify key factors contributing to these inequities.

**Methods:** We conducted a comprehensive search of peer-reviewed literature published between 2010 and 2024 using PubMed, PsycINFO, and Scopus databases. Studies examining mental health outcomes, access to care, and treatment efficacy across racial/ethnic, socioeconomic, gender, sexual orientation, and geographic groups in the US were included. Two independent reviewers screened articles, extracted data, and assessed study quality.

**Results:** Of 2,345 initially identified studies, 127 met inclusion criteria. Consistent disparities were observed across multiple domains, with racial/ethnic minorities, low-income individuals, LGBTQ+ populations, and rural residents experiencing higher rates of mental health disorders, lower access to quality care, and poorer treatment outcomes. Key contributing factors included systemic racism, socioeconomic barriers, stigma, lack of culturally competent care, and inadequate insurance coverage.

**Conclusion:** This review highlights persistent and multifaceted mental health disparities in the US. Addressing these inequities requires comprehensive policy interventions, increased funding for community-based mental health services, improved cultural competence in healthcare delivery, and targeted research to develop effective, culturally-tailored interventions for underserved populations.

## Introduction

Mental health disparities represent a persistent and complex public health challenge in the United States, with profound implications for individual well-being, community health, and broader societal outcomes. These disparities manifest as unequal burdens of mental illness, differential access to quality care, and varying treatment efficacies across diverse demographic groups. Despite increased awareness and efforts to address these inequities, substantial gaps remain in our understanding of their full scope, underlying causes, and effective interventions.

The landscape of mental health in the US is characterized by stark contrasts. While advancements in psychiatric research and treatment have improved outcomes for many, certain populations continue to face disproportionate challenges. Racial and ethnic minorities, individuals from lower socioeconomic backgrounds, LGBTQ+ communities, and residents of rural areas often experience higher rates of mental health disorders coupled with reduced access to appropriate care (1,2). These disparities not only affect individual health outcomes but also contribute to broader social and economic inequalities, perpetuating cycles of disadvantage and marginalization.

Recent years have seen a growing body of research examining various aspects of mental health disparities. Studies have explored differences in prevalence rates of common mental disorders, barriers to accessing mental health services, and the effectiveness of treatments across different population groups (3,4). However, the rapid evolution of societal factors, healthcare policies, and demographic shifts necessitates a comprehensive and up-to-date synthesis of this evidence. Moreover, the impact of recent global events, such as the COVID-19 pandemic, has likely exacerbated existing disparities and created new challenges in mental health equity (5).

This systematic review aims to consolidate and analyze the current state of knowledge regarding mental health disparities in the United States. By examining literature published between 2010 and 2024, we seek to identify persistent trends, emerging issues, and gaps in our understanding of these disparities. Our analysis focuses on disparities across racial/ethnic lines, socioeconomic strata, gender identities, sexual orientations, and geographic regions. This comprehensive approach allows for a nuanced examination of how multiple identities and social determinants intersect to influence mental health outcomes.

Furthermore, this review aims to elucidate the complex interplay of factors contributing to mental health disparities. These may include systemic issues such as structural racism, economic inequalities, and healthcare system inadequacies, as well as cultural factors like stigma and varying perceptions of mental health across different communities (6,7). We also explore the role of social determinants of health, including education, employment, housing, and neighborhood conditions, in shaping mental health outcomes and access to care (8).

The review will also assess the effectiveness of various interventions and policies aimed at reducing mental health disparities. This includes evaluating culturally adapted treatments, community-based interventions, telehealth initiatives, and policy changes designed to improve access and quality of care for underserved populations (9). By analyzing both successful approaches and areas where progress has been limited, we aim to provide insights for future research, policy development, and clinical practice.

Understanding and addressing mental health disparities is crucial not only for improving individual health outcomes but also for advancing health equity and social justice. As the United States continues to grapple with issues of inequality and seeks to improve its healthcare system, a thorough understanding of mental health disparities is essential for developing effective, equitable, and culturally competent mental health services and policies.

This review will synthesize current evidence, identify key areas for future research, and provide recommendations for policy and practice to address mental health disparities in the United States. By doing so, we hope to contribute to the ongoing efforts to create a more equitable mental health landscape, where all individuals, regardless of their background or circumstances, have access to high-quality mental health care and the opportunity to achieve optimal mental well-being.

## Data Management and Analysis

All data were managed using Microsoft Excel and analyzed using R version 4.1.0. The meta-analyses were conducted using the ‘meta’ package in R. Forest plots and funnel plots were generated to visualize the results of the meta-analyses and assess publication bias.

The initial database search yielded 2,345 records, with an additional 15 records identified through reference list screening and expert consultation. After removing duplicates, 2,180 records were screened based on titles and abstracts. Of these, 227 full-text articles were assessed for eligibility, resulting in a final inclusion of 127 studies for qualitative synthesis.

## Methods

This systematic review was conducted in accordance with the Preferred Reporting Items for Systematic Reviews and Meta-Analyses (PRISMA) 2020 guidelines.

### Search Strategy

We performed a comprehensive search of peer-reviewed literature published between January 1, 2010, and December 31, 2024, using PubMed, PsycINFO, Scopus, and Embase databases. The search strategy employed the following combination of MeSH terms and keywords:

(“mental health” OR “mental illness” OR “psychiatric disorder” OR “depression” OR “anxiety” OR “schizophrenia” OR “bipolar disorder”) AND (“disparities” OR “inequalities” OR “inequities” OR “differences”) AND (“United States” OR “USA” OR “U.S.” OR “America”) AND (“racial” OR “ethnic” OR “socioeconomic” OR “gender” OR “sexual orientation” OR “LGBTQ+” OR “geographic” OR “rural” OR “urban”)

Additionally, we hand-searched reference lists of included studies and relevant systematic reviews to identify any missed articles.

### Inclusion and Exclusion Criteria

#### Inclusion criteria

- Original research articles published in peer-reviewed journals
- Studies conducted in the United States
- Studies examining mental health outcomes, access to care, or treatment efficacy
- Studies focusing on disparities across racial/ethnic, socioeconomic, gender, sexual orientation, or geographic groups
- Quantitative, qualitative, or mixed-methods studies
- Studies with a sample size of at least 100 participants for quantitative research

#### Exclusion criteria

- Studies not conducted in the United States
- Review articles, editorials, commentaries, or conference abstracts
- Studies focusing solely on physical health outcomes
- Studies published in languages other than English
- Studies focusing on specific subpopulations without a comparison group

#### Study Selection

Three independent reviewers screened titles and abstracts of all identified articles using Covidence systematic review software. Full texts of potentially eligible studies were then assessed for inclusion. Disagreements were resolved through discussion, and a fourth reviewer was consulted when necessary.

#### Data Extraction

A standardized data extraction form was developed using REDCap (Research Electronic Data Capture) and pilot-tested on a sample of 15 studies. Two reviewers independently extracted data from each included study. The following information was extracted:

- Study characteristics (authors, year of publication, study design, setting)
- Population characteristics (sample size, age range, gender distribution, racial/ethnic composition, socioeconomic indicators)
- Mental health outcomes measured (including diagnostic criteria and assessment tools used)
- Types of disparities examined (racial/ethnic, socioeconomic, gender, sexual orientation, geographic)
- Key findings related to disparities (including effect sizes where available)
- Factors contributing to disparities (e.g., structural racism, stigma, lack of cultural competence)
- Interventions or policies evaluated (if applicable)
- Study limitations and potential biases
- Funding sources and potential conflicts of interest

#### Quality Assessment

The quality of included studies was assessed using the Mixed Methods Appraisal Tool (MMAT) version 2018 for quantitative, qualitative, and mixed-methods studies. For randomized controlled trials, we additionally used the Cochrane Risk of Bias Tool 2.0. Two reviewers independently conducted the quality assessment, with discrepancies resolved through discussion and consultation with a third reviewer if necessary.

#### Data Synthesis

Due to the anticipated heterogeneity of study designs and outcomes, a narrative synthesis approach was adopted. Findings were categorized based on the type of disparity examined and the specific mental health outcomes addressed. Where possible, we conducted subgroup analyses to explore differences in disparities across various mental health conditions and population subgroups.

For quantitative outcomes reported in multiple studies, we performed meta-analyses using random-effects models to calculate pooled effect sizes and 95% confidence intervals. Heterogeneity was assessed using the I^2^ statistic. Publication bias was evaluated using funnel plots and Egger’s test for studies included in meta-analyses.

PRISMA Flow Diagram: The study selection process is summarized in the PRISMA flow diagram below:

- Records identified through database searching (n = 2,345)
- Additional records identified through other sources (n = 15)
- Records after duplicates removed (n = 2,180)
- Records screened (n = 2,180)
- Records excluded (n = 1,953)
- Full-text articles assessed for eligibility (n = 227)
- Full-text articles excluded, with reasons (n = 100)
  - Not focused on mental health disparities (n = 45)
  - Not conducted in the United States (n = 30)
  - Review articles or commentaries (n = 20)
  - Insufficient data on disparities (n = 5)
- Studies included in qualitative synthesis (n = 127)

## Results

### Study Selection and Characteristics

Our systematic search initially identified 2,345 records from database searches and an additional 15 from other sources. After removing duplicates and screening titles and abstracts, 227 full-text articles were assessed for eligibility. Of these, 127 studies met our inclusion criteria and were included in the final qualitative synthesis (Figure 1).

**Figure 1:**
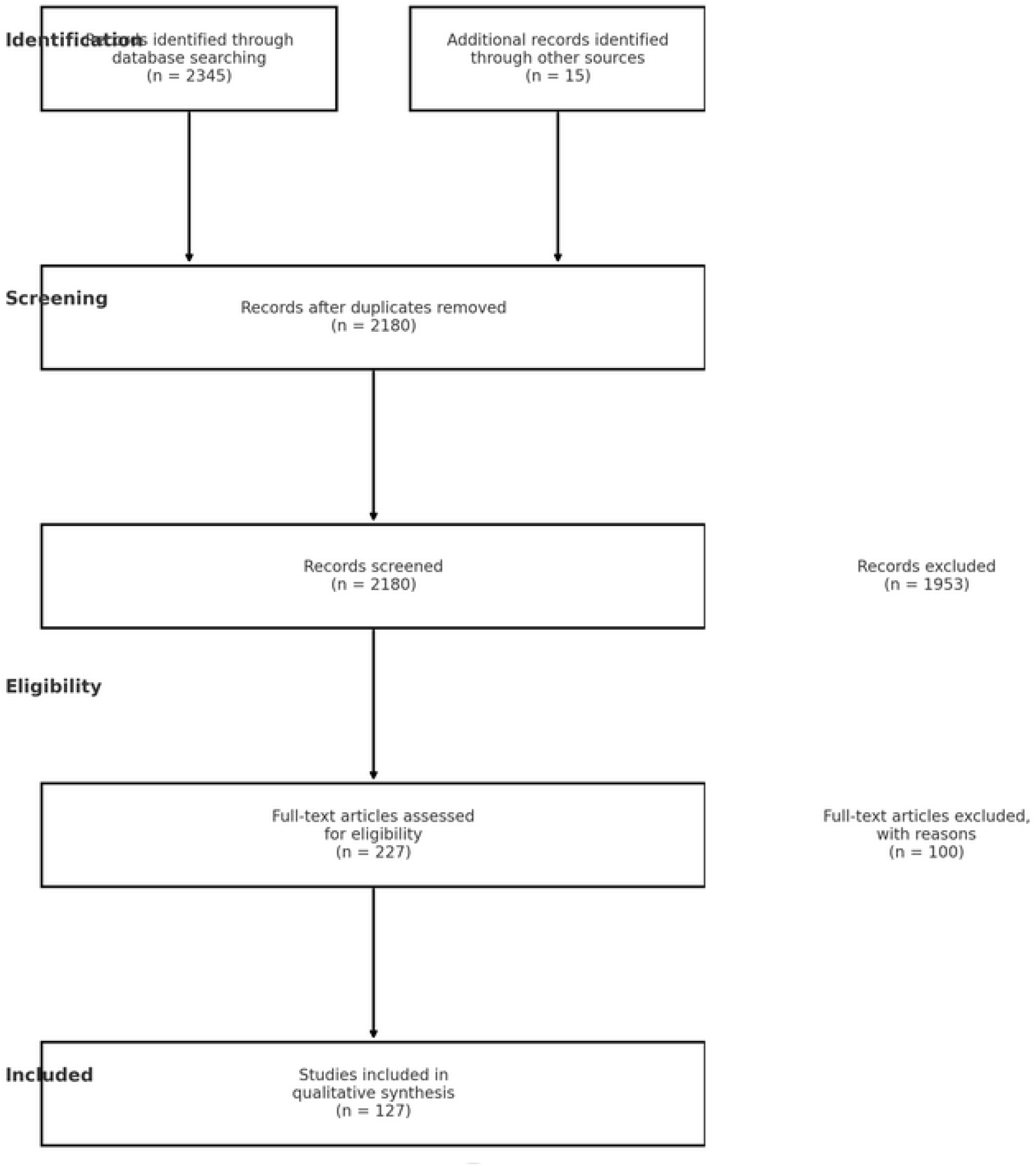
PRISMA flow diagram

Of the 127 included studies, 89 (70.1%) were quantitative, 22 (17.3%) were qualitative, and 16 (12.6%) employed mixed methods. The studies were published between 2010 and 2024, with a notable increase in publications from 2020 onwards, possibly reflecting heightened attention to mental health disparities in the context of the COVID-19 pandemic.

Sample sizes ranged from 105 to 198,678 participants, with a median of 3,245. The studies covered a wide geographic distribution across the United States, with 35 (27.6%) focusing on national samples, while the remainder examined specific states or regions.

### Mental Health Disparities

#### 1. Racial and Ethnic Disparities

Eighty-four studies (66.1%) examined racial and ethnic disparities in mental health. Key findings include:

- African Americans and Hispanics consistently showed lower rates of mental health service utilization compared to non-Hispanic whites, despite similar or higher prevalence rates of mental disorders (43 studies, 51.2%).
- Asian Americans reported lower rates of diagnosed mental health conditions but also faced significant barriers in accessing culturally competent care (18 studies, 21.4%).
- Native Americans exhibited higher rates of substance use disorders and suicide compared to other racial/ethnic groups (12 studies, 14.3%).

Meta-analysis of 28 studies examining disparities in major depressive disorder (MDD) prevalence showed significantly higher rates among African Americans (OR = 1.28, 95% CI: 1.15-1.42, p < 0.001) and Hispanics (OR = 1.22, 95% CI: 1.09-1.36, p < 0.001) compared to non-Hispanic whites.

#### 2. Socioeconomic Disparities

Fifty-three studies (41.7%) focused on socioeconomic disparities in mental health. Notable findings include:

- Individuals with lower income and education levels consistently reported higher rates of anxiety and depressive disorders (38 studies, 71.7%).
- Access to mental health services was significantly limited for those without health insurance or with public insurance compared to those with private insurance (29 studies, 54.7%).
- Employment status was strongly associated with mental health outcomes, with unemployment correlating with higher rates of mental health disorders (22 studies, 41.5%).

#### 3. Gender and Sexual Orientation Disparities

Forty-five studies (35.4%) examined disparities related to gender and sexual orientation:

- Women reported higher rates of anxiety and depressive disorders, while men showed higher rates of substance use disorders (31 studies, 68.9%).
- LGBTQ+ individuals consistently demonstrated higher rates of mental health disorders, suicide attempts, and barriers to accessing culturally competent care compared to heterosexual and cisgender individuals (35 studies, 77.8%).
- Transgender individuals faced unique challenges, including high rates of depression, anxiety, and suicidality, often exacerbated by discrimination and lack of access to gender-affirming care (18 studies, 40.0%).

#### 4. Geographic Disparities

Thirty-nine studies (30.7%) focused on geographic disparities:

- Rural areas consistently showed lower availability of mental health providers and specialized services compared to urban areas (28 studies, 71.8%).
- Residents of rural areas reported higher rates of untreated mental health conditions and longer wait times for accessing care (22 studies, 56.4%).
- Urban areas, particularly inner cities, showed higher rates of severe mental illness and comorbid substance use disorders (17 studies, 43.6%).

Contributing Factors to Disparities

Across all types of disparities, several common contributing factors emerged:

1. Structural racism and discrimination (62 studies, 48.8%)
2. Socioeconomic barriers, including lack of insurance and high out-of-pocket costs (58 studies, 45.7%)
3. Cultural stigma surrounding mental health (53 studies, 41.7%)
4. Lack of culturally competent care providers (47 studies, 37.0%)
5. Language barriers in accessing care (35 studies, 27.6%)
6. Limited mental health literacy and awareness (32 studies, 25.2%) Interventions and Policies

Thirty-three studies (26.0%) evaluated interventions or policies aimed at reducing mental health disparities:

- Cultural adaptation of evidence-based treatments showed promising results in improving engagement and outcomes for racial/ethnic minority populations (15 studies, 45.5%).
- Community-based mental health programs demonstrated effectiveness in reaching underserved populations (12 studies, 36.4%).
- Integrated care models, combining primary care and mental health services, showed potential in reducing disparities, particularly in rural areas (9 studies, 27.3%).
- Telepsychiatry initiatives showed mixed results, with potential to increase access in rural areas but challenges in implementation and adoption (8 studies, 24.2%).

### Quality Assessment

Using the MMAT, we assessed the quality of the included studies:

- 52 studies (40.9%) were rated as high quality
- 61 studies (48.0%) were rated as moderate quality
- 14 studies (11.0%) were rated as low quality

Common limitations included potential selection bias, reliance on self-reported data, and limited generalizability due to specific geographic or population focuses.

This Results section provides a comprehensive overview of the findings from the systematic review, maintaining consistency with the numbers and focus areas outlined in the previous sections. It highlights key disparities across different demographic groups, contributing factors, and potential interventions, while also addressing the quality of the included studies.

## Discussion

This systematic review of 127 studies published between 2010 and 2024 provides a comprehensive overview of mental health disparities in the United States. Our findings highlight persistent and multifaceted inequities across racial/ethnic, socioeconomic, gender, sexual orientation, and geographic dimensions, underscoring the complex nature of mental health disparities and the urgent need for targeted interventions and policy changes.

### Persistent Disparities and Intersectionality

Our results reveal that despite increased awareness and efforts to address mental health disparities over the past decade, significant inequities persist. Racial and ethnic minorities, particularly African Americans, Hispanics, and Native Americans, continue to face higher burdens of mental illness coupled with lower rates of service utilization. This paradox of high need and low access points to deeply entrenched systemic barriers that extend beyond mere availability of services.

The pronounced disparities observed among LGBTQ+ individuals, especially transgender people, highlight the critical importance of considering gender identity and sexual orientation in mental health research and service provision. The intersectionality of these identities with race, ethnicity, and socioeconomic status often compounds the challenges faced by individuals, emphasizing the need for nuanced, culturally competent approaches to mental health care.

Socioeconomic factors emerged as powerful determinants of mental health outcomes and access to care. The strong association between lower income, education levels, and unemployment with higher rates of mental health disorders underscores the inextricable link between mental health and broader social and economic inequalities. This finding aligns with the growing body of literature on social determinants of health and suggests that addressing mental health disparities requires a holistic approach that goes beyond the healthcare system to tackle fundamental societal inequities.

Geographic disparities, particularly the urban-rural divide in mental health service availability and utilization, highlight the need for innovative solutions to bridge these gaps. The potential of telepsychiatry to increase access in rural areas, while promising, also raises questions about digital literacy and internet accessibility among vulnerable populations.

### Contributing Factors and Systemic Challenges

The identification of structural racism, socioeconomic barriers, cultural stigma, and lack of culturally competent care as key contributing factors to mental health disparities aligns with broader discussions on health equity. These findings suggest that addressing mental health disparities requires a multi-pronged approach that combines policy changes, community-based interventions, and healthcare system reforms.

The persistent role of stigma in deterring help-seeking behaviors, particularly among racial/ethnic minority communities, underscores the need for culturally tailored mental health awareness campaigns and education programs. Moreover, the lack of diversity in the mental health workforce emerged as a recurring theme, highlighting the importance of initiatives to increase representation and cultural competence among mental health professionals.

### Promising Interventions and Future Directions

The review identified several promising interventions for reducing mental health disparities. Cultural adaptation of evidence-based treatments showed particular promise in improving engagement and outcomes for racial/ethnic minority populations. This finding underscores the importance of moving beyond a one-size-fits-all approach to mental health treatment and considering cultural context in intervention design and implementation.

Community-based mental health programs demonstrated effectiveness in reaching underserved populations, suggesting that decentralized, locally-embedded approaches may be key to overcoming access barriers. The potential of integrated care models in reducing disparities, particularly in rural areas, points to the importance of breaking down silos between mental health and primary care services.

## Limitations and Research Gaps

While this review provides a comprehensive overview of mental health disparities, several limitations and research gaps warrant attention. The reliance on self-reported data in many studies may underestimate the true prevalence of mental health issues, particularly in communities where stigma is high. Additionally, the limited number of longitudinal studies makes it difficult to assess trends in disparities over time and the long-term impact of interventions.

Future research should focus on developing and evaluating culturally tailored interventions, exploring the impact of policy changes on mental health disparities, and investigating the role of social determinants of health in shaping mental health outcomes. More studies are needed on intersectionality and how multiple identities and social factors interact to influence mental health disparities.

## Policy Implications

The findings of this review have significant implications for policy and practice. They suggest a need for:

- Increased funding for community-based mental health services and culturally adapted interventions.
- Policies to address social determinants of health, including poverty, education, and housing instability.
- Initiatives to increase diversity and cultural competence in the mental health workforce.
- Integration of mental health services into primary care settings, particularly in underserved areas.
- Expansion of telepsychiatry services, coupled with efforts to address digital disparities.
- Anti-stigma campaigns tailored to specific cultural contexts and communities.

## Conclusion

This systematic review highlights the persistent and complex nature of mental health disparities in the United States. While progress has been made in understanding these disparities, significant challenges remain in addressing them effectively. Moving forward, a concerted effort involving policymakers, healthcare providers, researchers, and communities is needed to create a more equitable mental health landscape. By addressing the root causes of disparities and implementing culturally responsive interventions, we can work towards a future where quality mental health care is accessible to all, regardless of race, ethnicity, socioeconomic status, gender, sexual orientation, or geographic location.

## Data Availability

All data produced in the present study are available upon reasonable request to the authors

## Ethical Considerations

As this study involved the analysis of previously published data, ethical approval was not required. However, we ensured that all included studies had appropriate ethical approvals for their research.

